# Potential Relationship Between eGFR_cystatin C_/eGFR_creatinine_-ratio and Glomerular Basement Membrane Thickness in Diabetic Kidney Disease

**DOI:** 10.1101/2020.12.16.20248179

**Authors:** Carl M. Öberg, Martin Lindström, Anders Grubb, Anders Christensson

**Affiliations:** Department of Clinical Sciences Lund, Lund University, Department of Nephrology, Skåne University Hospital, Lund, Sweden; Department of Laboratory Medicine Malmö, Lund University, Center for Molecular Pathology, Skåne University Hospital, Malmö, Sweden; Department of Laboratory Medicine, Lund University, Department of Clinical Chemistry, Skåne University Hospital, Lund, Sweden; Department of Clinical Sciences Malmö, Lund University, Skåne University Hospital, Department of Nephrology, Malmö, Sweden

**Keywords:** diabetic kidney disease, estimated GFR, two-pore model, glomerular barrier thickness, ratio eGFR_cystatinC_/eGFR_creatinine_

## Abstract

**Background:** Diabetic nephropathy (DN) is a leading cause of end stage renal disease and renal replacement therapy worldwide. A pathophysiological hallmark of DN is glomerular basal membrane (GBM) thickening, whereas this feature is absent in minimal change disease. Theoretically, a thicker GBM will impede the diffusion of middle-molecules such as cystatin C, potentially leading to a lower estimated GFR (eGFR) from cystatin C compared to that of creatinine. Here we test the hypothesis that thickening of the glomerular filter leads to an increased diffusion length, and lower clearance, of cystatin C.

**Methods:** Twenty-nine patients with a kidney biopsy diagnosis of either DN (n=17) or minimal change disease (MCD) (n=12) were retrospectively included in the study. GBM thickness was measured at 20 separate locations in the biopsy specimen and plasma levels of cystatin C and creatinine were retrieved from health records. A modified two-pore model was used to simulate the effects of a thicker GBM on glomerular water and solute transport.

**Results:** The mean age of the patients was 52 years, and 38% were women. The mean eGFR_cystatin C_/eGFR_creatinine_-ratio was 74% in DKD compared to 98% in MCD (P < 0.001). Average GBM thickness was strongly inversely correlated to the eGFR_cystatin C_/eGFR_creatinine_-ratio (Pearson’s r=-0.61, P < 0.01). Two-pore modeling predicted a eGFR_cystatin C_/eGFR_creatinine_-ratio of 78% in DN.

**Conclusions:** We provide clinical and theoretical evidence suggesting that thickening of the glomerular filter, increasing the diffusion length of cystatin C, lowers the eGFR_cystatin C_/eGFR_creatinine_-ratio in DN.

**Significance statement:** Increased thickness of the glomerular basement membrane (GBM) is an early structural abnormality in diabetes, and has been identified as an independent risk factor for progression to diabetic nephropathy (DN). Increased GBM thickness will increase the diffusion length of mid-sized molecules like cystatin C across the renal filter, potentially leading to increased plasma concentrations and lower estimated GFR from cystatin C. The authors show theoretically that estimated GFR from cystatin C (eGFR_cystatin C_) is lower than that of creatinine (eGFR_creatinine_) in DN to a degree directly depending on GBM thickness. In line with theory, they provide clinical data showing a lower eGFR_cystatin C_ in diabetic patients with the eGFR_cystatin C_/eGFR_creatinine_-ratio being inversely correlated to GBM thickness assessed from electron micrographs.

## Introduction

The number of patients with diabetes and chronic kidney disease (CKD) is rapidly increasing worldwide ^1^. Advanced diabetic kidney disease is considered an irrecoverable condition, and identifying diagnostic methods and applying early interventions are crucial to reduce the global burden of end stage renal disease.

Cystatin C has emerged as a marker for noninvasive estimation of GFR ^2^ ever since it was noted that its reciprocal concentration correlated closely with GFR ^3^. Compared to creatinine, it appears to be especially well suited for the detection of early impairments in renal function ^4, 5^. Cystatin C has a rather high molecular weight (Mw) of 13.3 kDa and a Stokes-Einstein radius of ∼1.8 nm compared to that of smaller endogenous markers such as creatinine (MW 0.1 kDa and SE-radius 0.3 nm). Nonetheless, cystatin C is regarded as a better predictor of adverse outcomes ^6, 7^ as well as a better marker for GFR in combination with creatinine ^8, 9^ compared to creatinine alone.

The glomerular clearance of a small molecule like creatinine should very closely match that of the permeate flux (i.e. the glomerular filtration rate) over the glomerular filtration barrier (GFB) whereas the clearance of larger molecules will be increasingly dependent on the size of the selective elements in the GFB (i.e. the functional small pore radius). In fact, while most of the plasma content of cystatin C and other small plasma proteins are catabolized in the renal tubules ^10^, cystatin C is not freely filtered across the GFB (sieving coefficient 0.84). Being a middle-sized molecule it likely has a glomerular fractional clearance similar to that of β_2_-microglobulin ^11^ and other small plasma proteins ^3^ (or 26 å dextran ^12^).

Shrunken pore syndrome (SPS) is defined as a difference in estimated GFR from cystatin C vs. creatinine (eGFR_Cystatin C_ < 60% eGFR_Creatinine_ in the absence of non-renal influence on the levels of cystatin C or creatinine ^13^), and has been identified as a strong independent risk factor for mortality in patients undergoing elective coronary artery bypass grafting ^9^ and chronic kidney disease. This effect was seen also regardless of eGFR level ^14^. The suggested mechanism for SPS is a shrinking of the glomerular pore size leading to an increased plasma concentration of small plasma proteins like cystatin C, β_2_-microglobulin, beta-trace protein and retinol binding protein ^13, 15^. Small plasma proteins like cystatin C are cleared from the body by glomerular filtration and degraded by renal tubular catabolism whereas only smaller part of large proteins are cleared in the kidneys ^16, 17^. Thus, when the glomerular filtration rate is decreased due to renal disease, plasma concentrations of small plasma solutes will increase ^15^ which is the basis of noninvasive GFR estimation.

A decreased pore size is not the only possible mechanism leading to SPS and/or reduced estimated GFR from cystatin C *vs*. creatinine, but could potentially result also from a thicker glomerular barrier leading to an increased diffusion length for middle molecules like cystatin C. One of the earliest structural alterations in diabetic kidney disease (DKD), preceding microproteinuria, is thickening of the GBM ^18^ whereas this condition is absent in minimal change disease (MCD). The aim of the current study was to test the hypothesis that estimated GFR from cystatin C is lower compared to that of creatinine in DKD vs. MCD, and that this difference is correlated to GBM thickness. To explore if the observed alterations can be explained theoretically, we used the heteroporous two-pore model by Deen and colleagues^19^ adapted to human physiological conditions. Due to the limited experimental information available on the hemodynamic conditions in the human glomerulus, some physiological parameters had to be estimated based on previous modeling by Oken ^20^ and Navar *et al* ^21^.

## Methods

### Design, Setting, and Participants

All patients referred for a kidney biopsy in Skåne, southern part of Sweden, are registered in a local data repository called the *Örestadsregistret (Örestad Registry)*. We assessed 579 entries in the Örestad Registry between August 2013 and August 2017. Inclusion criteria were biopsy diagnosis of diabetic kidney disease or minimal change disease and estimated GFR measurement using both cystatin C and creatinine within 2 weeks of the time of biopsy. All patients were assessed for medication including angiotensin-converting enzyme inhibitors/angiotensin-receptor blockers and corticosteroids. Exclusion criteria were signs of other pathologies in the biopsy specimen, e.g. Ig A nephropathy or hypertensive nephrosclerosis. We identified 12 patients with minimal change disease and 17 patients with DKD who fulfilled the inclusion criteria and none of the exclusion criteria. The Research Ethics Board at Lund University approved the study (Dnr 2017/568). Patient data and samples were treated anonymously in all statistical analyses. This study is in accordance with the recommendations outlined in the “STAndards for the Reporting of Diagnostic accuracy studies” (STARD) statement ^22^ (Supplemental File 1).

### Laboratory measurements for renal function

Plasma concentrations of creatinine and cystatin C were collected from health records. Cystatin C was determined by a particle-enhanced immunoturbidimetric method using a reference material traceable to the international cystatin C calibrator ^23^. Creatinine was determined by an enzymatic colorimetric assay using a calibrator traceable to primary reference material with values assigned by isotope dilution mass spectrometry ^24^. GFR was estimated (eGFR) from the LMrev formula based on creatinine ^24^ and the CAPA formula based on cystatin C ^23^.

### A modified two-pore model to simulate the effects of a thicker GBM on glomerular water and solute transport

Three pathophysiological changes will affect glomerular solute and water transport in DKD: 1. GBM hypertrophy, 2. glomerular hypertension and 3. hyperfiltration (single nephron). We simulated these changes by 1. lowering the filtration coefficient (LpA) and solute diffusion capacity (half for twice the thickness and so on), 2. increasing the glomerular hydraulic pressure gradient to achieve 3. a single nephron GFR (SNGFR) of 90 nl/min. If the glomerular filtration coefficient, hydraulic pressure gradient, single nephron GFR, and (afferent) oncotic pressure are known, then the single nephron plasma flow profile Q(y) and the plasma protein concentration C_pr_(y) as functions of the relative position y (position/total length) along the glomerular capillary, can be determined by use of the conservation of mass equations

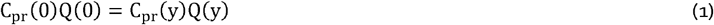

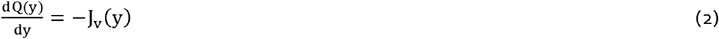

Re-arranging these equations gives the well-known ^19, 25, 26^ non-linear ordinary differential equation

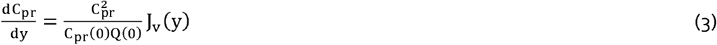

Above, J_v_(y) is the local filtration flux ^26^ which is equal to the local Starling pressure (difference between hydraulic and oncotic pressure gradients) multiplied by the ultrafiltration coefficient (L_p_A)

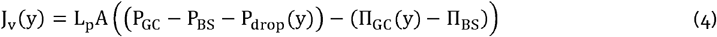

The colloid osmotic pressure in the glomerular capillary (Π_**GC**_) was calculated using the Landis-Pappenheimer equation, 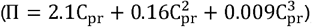, assuming a total plasma protein concentration of 6.7 g/dL corresponding to a plasma colloid osmotic pressure of ∼24 mmHg. In the human glomerulus, the capsular hydrostatic pressure (P_BS_) has been estimated to be between 15-20 mmHg ^20, 21^ and the capsular oncotic pressure (Π_**BS**_) 0 mmHg. In order to achieve an effective filtration pressure (EFP) in the normal state of ∼10 mmHg, we assumed a glomerular hydraulic pressure gradient of 40 mmHg (cf. ^21^ and ^20^), implying a glomerular capillary pressure (P_GC_) of 55-60 mmHg. The pressure drop along the glomerular capillary network was set to 1 mmHg ^19^. The renal plasma flow (RPF or Q(0)) was determined with the gradient descent algorithm described in ^25^.

Actual measurements of glomerular sieving coefficients for small proteins like cystatin C across the human glomerular filter are scarce in the literature. Perhaps the most reliable estimations are those obtained in children with Fanconi syndrome ^17, 27^, although these sieving coefficients may be slightly low due to residual tubular reabsorption ^27^. Based on the sieving data in ^17^ and previous results from the rat glomerular filtration barrier we assumed a small pore radius (r_S_) of 3.7 nm and a large pore radius (r_L_) of 11.0 nm (cf. ^11^). Again, if assuming conservation of mass (see also ^25^), the change in solute (protein) concentration C_i_(y) due to glomerular filtration (at position y along the glomerular capillary) is given by

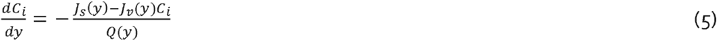

where the local solute flux of a solute, J_s_(y), is calculated from

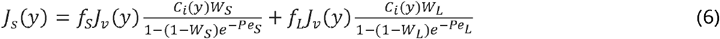

where C_i_(y) is the solute concentration at position *y* along the glomerular capillary; f_S_ and f_L_ are the fractional volume flows; W_S_ and W_L_ are the convective hindrance factors ^28^, across the small- and the large pores respectively. Péclet numbers (Pe_S_ and Pe_L_) are calculated from

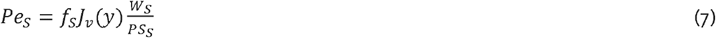

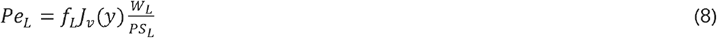

Solute diffusion capacities PS_S_ and PS_L_ (cf. eqn 8 and 9 in ^29^) were calculated from

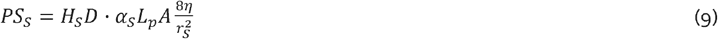

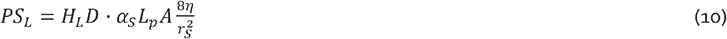

where H_S_ and H_L_ are the diffusive hindrance factors ^28^, D is the free diffusion coefficient calculated from the Stokes-Einstein radius of the solute protein, η is the viscosity of the permeate (0.7 mPa·s), α_**S**_ and α_**L**_ denote the fractional hydraulic conductance of the small and large pore system, respectively (for a discussion on the differences between α_**S/L**_ and f_S/L_ and their calculation, see ^30^). In the current article the f_L_ parameter was set to ∼1·10^-4^ (similar to that obtained in humans using Ficoll ^31^) which gives a α_**L**_ of ∼3·10^-5^. We assumed a steady state plasma clearance of cystatin C (and creatinine), occurring chiefly via glomerular filtration:

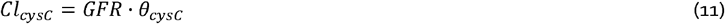

where θ_cysC_ is the glomerular sieving coefficient of cystatin C. A small pore system with a 3.7 nm pore radius implies a sieving coefficient for cystatin C of ∼0.87. For an adult male having a GFR of 120 mL/min, this translates to a cystatin C concentration of about 0.69 mg/L.

### Measurement of glomerular basal membrane (GBM)

Core biopsies were first put in 4% formaldehyde and thereafter embedded in paraffin. Cases were sectioned in two levels, each 1,5 um thick, and stained for HE, ABPAS, TRI, EVG, PASM and AK and each case were re-evaluated by light microscopy. Samples for electron microscopy (EM) were put in 4% formaldehyde and embedded in (plastic) and thereafter sectioned in 60 nm thick sections. Transmission electron microscopy was performed for each case with a transmission microscope (TECNAI FEJ Company) as well as photo documented in JPEG-format by a digital camera (VELETA). One glomerulus per case with at least one full segment was examined with electron microscopy. Each case was evaluated regarding basal membranes, foot processes and mesangium and were found to correspond to the diagnosis of diabetic nephropathy or minimal change nephropathy. Each case showed homogenous thickness of basal membranes of segments without any focal irregularities or deposits. For each glomeruli 20 measurements points were taken in a representative loop. GBM thickness was assessed through ultrastructural morphometry on enlarged electron micrographs as the arithmetic mean of 20 orthogonal intercepts across the GBM. Electron micrographs were available in 8 patients with MCD and in 11 patients with DKD.

### Statistical methods

Values are presented as average (IQR) unless otherwise specified. Differences between groups were assessed using a Wilcoxon signed rank test. Monte Carlo based power analysis (10,000 simulations) was conducted, applying Wilcoxon tests between fixed unequal sample sizes of 17 and 12 patients, showing that the study had 89% power to detect a true difference of 0.2 in eGFR_cystatin C_/eGFR_creatinine_-ratio assuming a SD of 0.16 (Cohen’s *d* = 1.25). Moreover, a sample size of 19 patients had 87% power to detect a true correlation coefficient of 0.6 or better between GBM-thickness and eGFR_cystatin C_/eGFR_creatinine_-ratio, at a significance level of 0.05. A post-hoc ANCOVA was applied using the model [ratio ∼ eGFR + diagnosis] using type III SS. We assumed an α-level of 0.05 and a β-level of 0.20 unless otherwise specified. P-values are denoted: * for P < 0.05, ** for P < 0.01 and *** for P > 0.001. All statistical calculations were performed using R for macOS (R Foundation for Statistical Computing, version 3.5.1)

## Results

### Reduced Ratio of eGFR_cystatin C_/eGFR_creatinine_ in Diabetic Kidney Disease vs. Minimal Change Disease

We first assessed estimated GFR (eGFR) from cystatin C and creatinine in two groups of patients with a biopsy diagnosis of either diabetic kidney disease (DKD) (n=17) or minimal change disease (MCD) (n=12). Baseline characteristics and eGFR estimations are shown in Table 1. Median age of the patients was 52 (41-64) years at the time of biopsy with no difference between the groups, and 38% were female. Plotted in Figure 1 A is the ratio eGFR_cystatin C_/eGFR_creatinine_ which was lower (*p*=0.0004, W=26) in DKD patients compared with MCD patients. Furthermore, 4 patients with DKD also fulfilled the criteria for shrunken pore syndrome, defined as eGFR_cystatin C_ < 60% eGFR_creatinine_ whereas none of the MCD patients fulfilled this criteria (the lowest ratio in the MCD group was ∼76%). Average estimated GFR (eGFR), calculated as the arithmetic average of eGFR_cystatin C_ and eGFR_creatinine_, was different in the groups, which is expected since GFR is usually much less affected in MCD (Table 1). To investigate if reduced eGFR was in itself correlated to a reduced ratio, we performed a *post-hoc* analysis of co-variance showing no significant main effect of eGFR on the eGFR_cystatin C_/eGFR_creatinine_-ratio (*F*_*1,26*_=0.2, *p*=0.65).

**Table 1.**
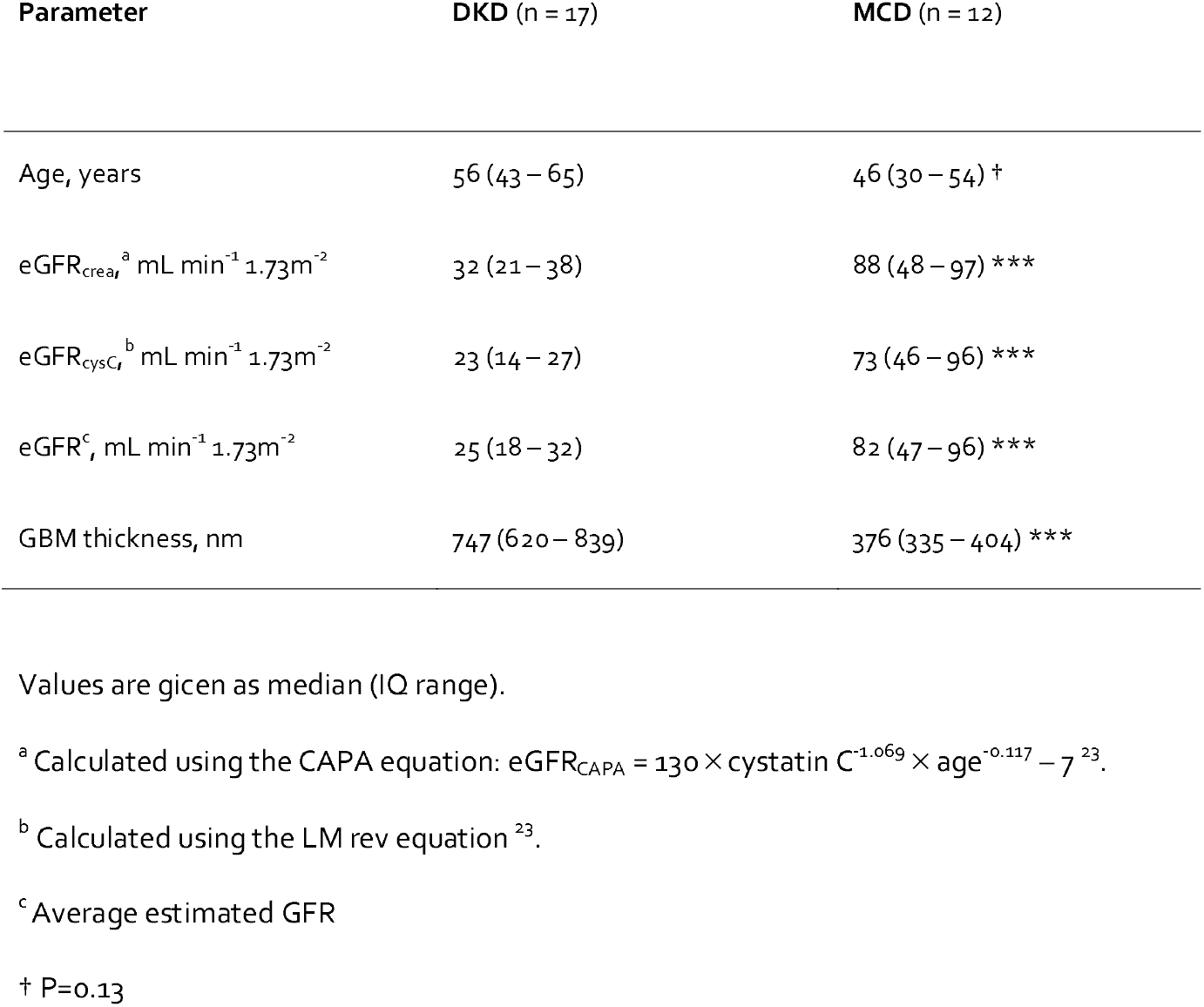
Clinical GFR estimations

**Figure 1.**
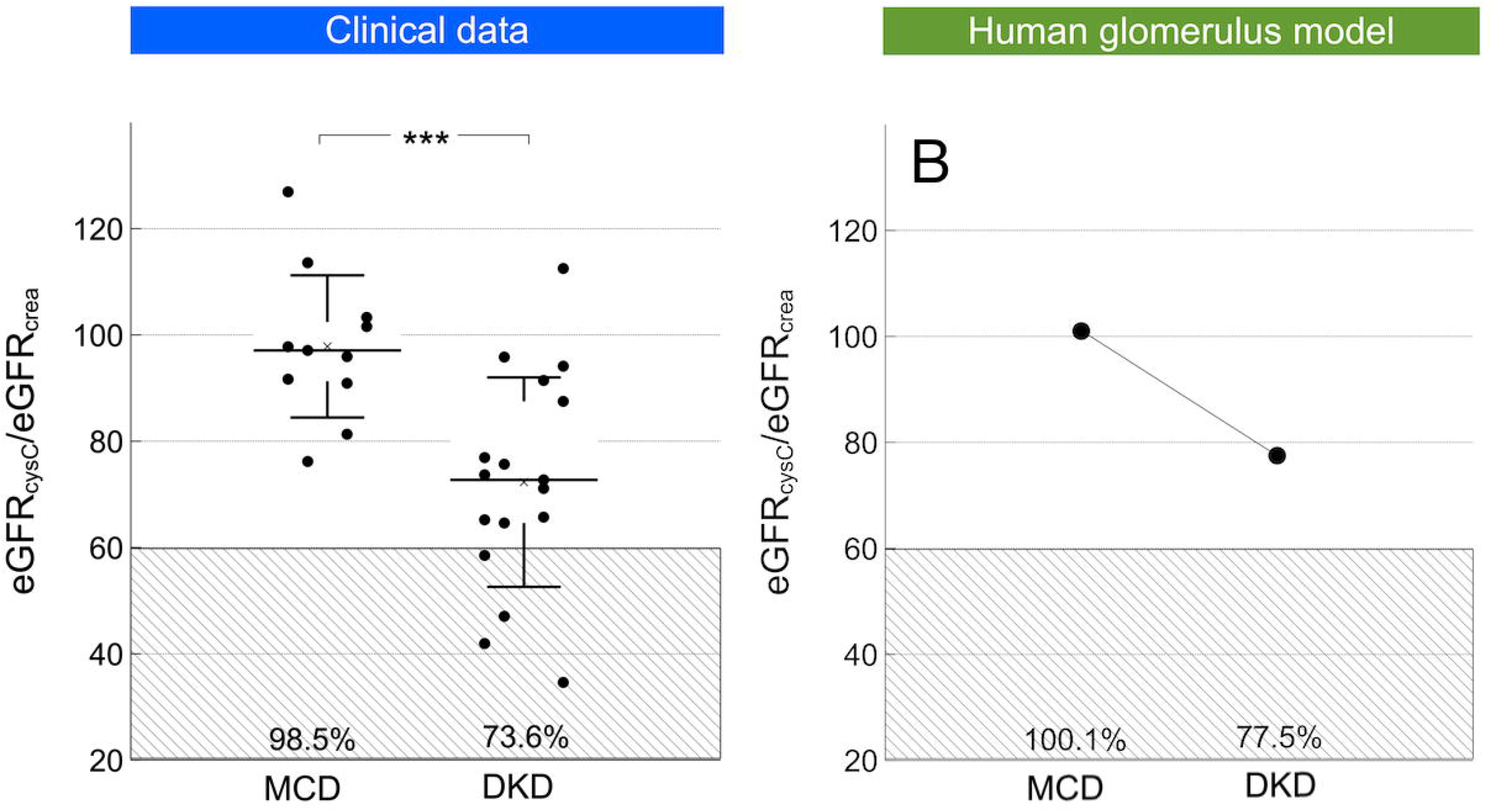
*A:* Estimated GFR from cystatin C and creatinine ratio in patients with a biopsy diagnosis of either diabetic kidney disease (DKD; n=17) or minimal change disease (MCD; n=12). *B:* Estimated GFR from cystatin C and creatinine in a theoretical model of the human glomerular filtration barrier. In silico simulations were performed either assuming a normal thickness of the glomerular basement membrane (GBM; 300 nm) or a hypertrophic GBM being 3 times thicker (900 nm). It was assumed that GFR remained constant as a result of tubulo-glomerular feedback, thus implying a three times higher effective filtration pressure across the glomerulus in the hypertrophic condition.

### The eGFR_cystatin C_/eGFR_creatinine_-ratio is strongly correlated to GBM thickness

Linear regression was performed to investigate the theoretically predicted relationship between the eGFR_cystatin C_/eGFR_creatinine_-ratio and GBM thickness (nm). The scatterplot (Figure 2) indeed suggested a negative linear relationship between the two, which was confirmed with a Pearson’s correlation coefficient of -0.61. Simple linear regression showed a significant relationship between eGFR_cystatin C_/eGFR_creatinine_ -ratio and GBM thickness (nm) (*p* < 0.01). The slope coefficient for mean GBM-thickness was -0.04 % nm^−1^ so the eGFR_cystatin C_/eGFR_creatinine_ -ratio was apparently reduced by ∼4% for every 100 nm increased thickness. As shown in Figure 2 B, the plot of residuals *vs*. the independent parameter indicated that the residuals were approximately normally distributed. To explore whether body composition and muscle mass affect the ratio eGFR_cystatin C_/eGFR_creatinine_ we used linear correlation between weight vs eGFR_cystatin C_/eGFR_creatinine_ -ratio and body mass index (BMI) vs eGFR_cystatin C_/eGFR_creatinine_ -ratio, but found no significant association.

**Figure 2.**
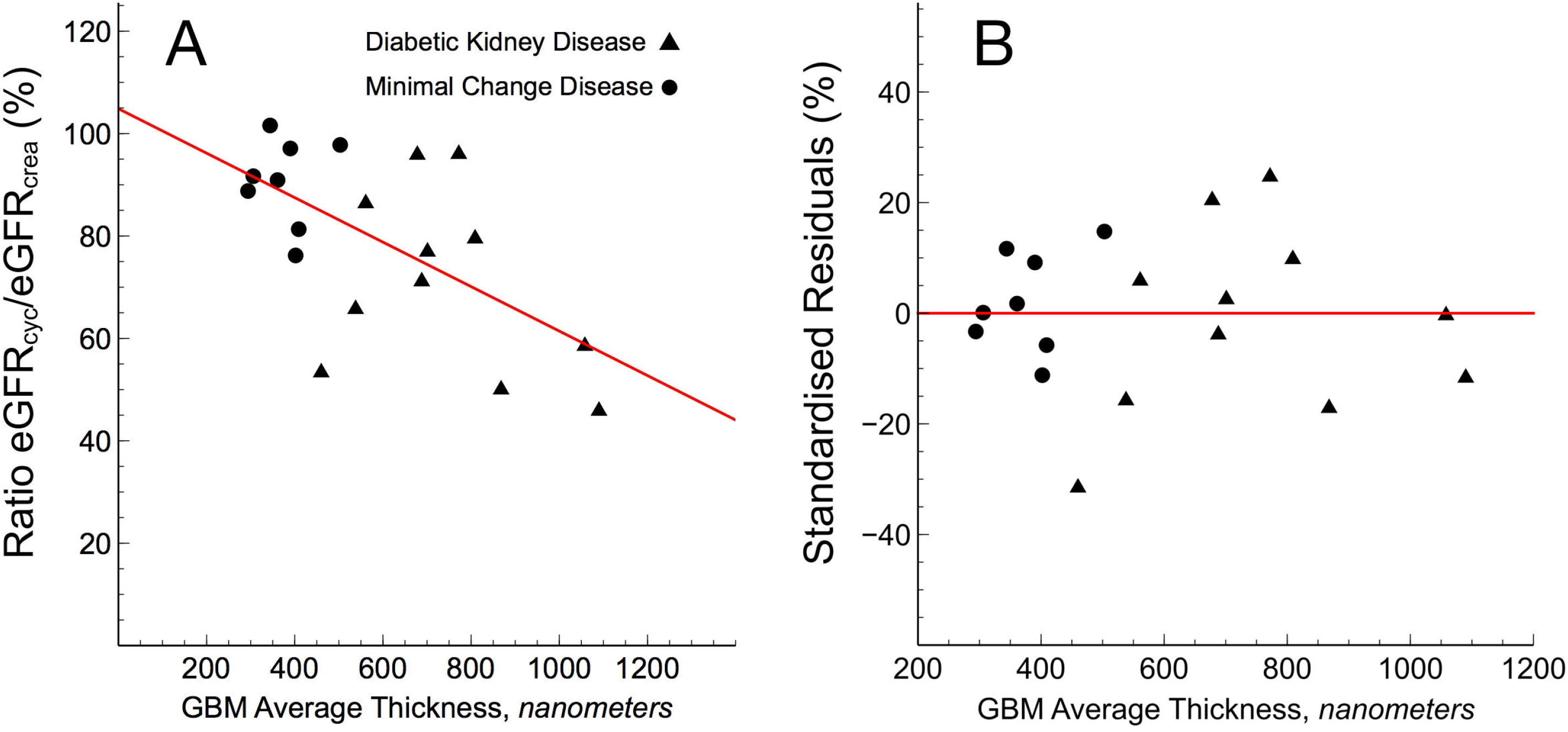
*A*: Estimated GFR from cystatin C and creatinine (eGFR_cystatin C_/eGFR_creatinine_) ratio versus glomerular basement membrane thickness as assessed via electron micrographs from kidney biopsies. *B*: Difference between modeled and measured eGFR_cystatin C_/eGFR_creatinine_-ratio as a function of glomerular basement membrane thickness.

### Reduced Ratio of eGFR_cystatin C_/eGFR_creatinine_ is theoretically dependent on GBM thickness

Pathophysiological hallmarks in the development of diabetic glomerulosclerosis are glomerular hypertrophy, hypertension and hyperfiltration; processes that were here modeled *in silico* by increasing the thickness of the GBM and the effective filtration pressure to result in a SNGFR of 90 nL min^-1^. We first increased glomerular pressure linearly with decreasing GFB conductance (+10 mmHg for twice the thickness, +20 mmHg for triple and so on). This had the effect of reducing the eGFR_cystatin C_/eGFR_creatinine_ ratio. Glomerular hypertension and GBM hypertrophy evoked marked reductions in the clearance of cystatin C (Figure 1 B & Figure 3). Plotted in Figure 1 B are the simulated ratios of eGFR_cystatin C_/eGFR_creatinine_ in a normal glomerulus having a glomerular basement membrane thickness of 300 nm and an effective filtration pressure (EFP) of 10 mmHg *versus* a “diabetic” glomerulus having a glomerular basement membrane thickness of 900 nm and an effective filtration pressure (EFP) of 30 mmHg. According to the current model, the impact of differently sized solutes is heterogeneous. Shown in Figure 3 are simulated glomerular sieving coefficients vs. the Stokes-Einstein radii of the solute proteins for three different scenarios. The solid line represents a normal healthy subject having a glomerular basement membrane thickness of ∼300 nm and an effective filtration pressure (EFP) of 10 mmHg. Also plotted are experimentally measured sieving coefficients for β_2_-microglobulin ^17^ (black square) and myoglobin ^11^ (black triangle). The simulated sieving coefficient of cystatin C is also shown (black hexagon). The sieving coefficient for cystatin C is, under normal conditions ∼0.84 corresponding to a glomerular clearance of ∼5/6 of GFR, i.e. Cl_CC_ ≈ 100 mL/min/1.73m^2^ for a relative GFR of 120 mL/min. For the scenarios with a thicker glomerular filter (dashed and dotted line), glomerular hypertension is assumed necessary to maintain the same GFR and the EFP will increase linearly with the thickness of the barrier, thus for a 600 nm barrier (dashed line) the EFP is 20 mmHg. This thickening of the barrier will also double the diffusion distance for cystatin C, effectively lowering its fractional clearance to 0.74 (Cl_CC_ ≈ 89 mL/min/1.73m^2^) for the 600 nm barrier and to 0.67 for the 900 nm barrier (Cl_CC_ ≈ 80 mL/min/1.73m^2^). Assuming a constant production of cystatin C, these latter scenarios will cause the serum concentration of cystatin C (and other small proteins) to increase so that the estimated GFR using e.g. the CAPA equation ^23^ will be lower than GFR. This effect is shown in Table 2. By contrast, the sieving coefficient of a 3.05 nm protein (here exemplified by neutral horseradish peroxidase nHRP ^11^; solid star) will be practically unaltered by the thickening of the renal filter.

**Table 2.** Glomerular hypertension and hypertrophy in a human glomerulus model

| Barrier thickness | 300 nm | 600 nm | 900 nm |
| --- | --- | --- | --- |
| GFR, mL/min/1.73 m<sup>2</sup> | 120 | 120 | 120 |
| Cystatin C, mg/L | 0.70 | 0.79 | 0.88 |
| eGFR<sub>CAPA</sub> † | 121 | 105 | 93 |

† eGFR<sub>CAPA</sub> = $130 \times \text{cystatin C}^{-1.069} \times \text{age}^{-0.117} - 7$

**Figure 3.**
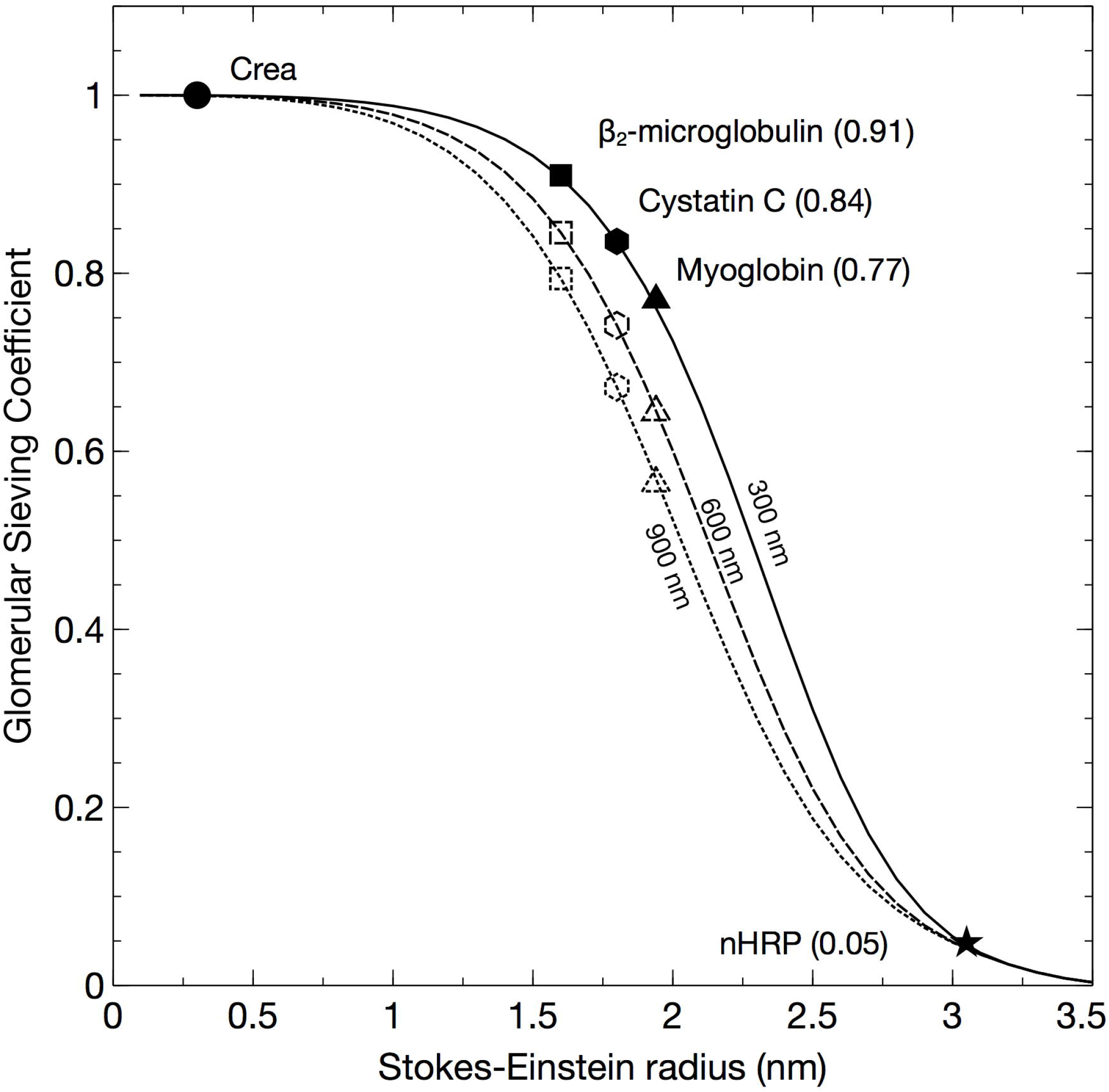
Glomerular sieving coefficients vs. the Stokes-Einstein radii of the solute proteins for three different thicknesses of the glomerular basement membrane 300 nm (and an effective filtration pressure (EFP) of 10 mmHg), 600 nm (EFP 20 mmHg), 900 nm (EFP 30 mmHg). Also shown are experimentally measured sieving coefficients for β_2_-microglobulin ^17^ (black square) and myoglobin ^11^ (black triangle). The sieving coefficient of a 3.05 nm protein (neutral horseradish peroxidase nHRP ^11^; solid star) will be practically unaltered by the thickening of the renal filter.

### Theoretical effects of actual ‘shrunken pores’ in the glomerular filter

In Figure 4, the modeled sieving coefficients vs. SE-radius are plotted for scenarios with shrunken pores, 3.4 nm (dashed line) and 3.2 nm (dotted line). Expectedly, shrinking the pore size causes the sieving curve to shift to the left. As can be seen, pore shrinking has similar effects to thickening of the GFB for smaller proteins. However, by contrast, it has marked effects for larger solutes having radii close to the pore radius (< 3.7 nm). For the 3.05 nm protein (nHRP, star) the sieving coefficient will be reduced by a factor ∼20 if the pores are shrunken to 3.2 nm. The effects of the different scenarios are demonstrated in Figure 5, where the ratio of the pathological vs. healthy scenarios in Figure 3 and 4 are plotted. Thus, an ideal solute size to detect GFB thickening would be approximately 2.5 nm whereas solutes exceeding 3.0 nm would be essentially unaffected under the current assumptions of an unchanged GFR, but the same effect will be present during conditions with a lower GFR. Actually, for slightly decreased GFRs, CrCl will typically overestimate the actual GFR contributing to SPS.

**Figure 4.**
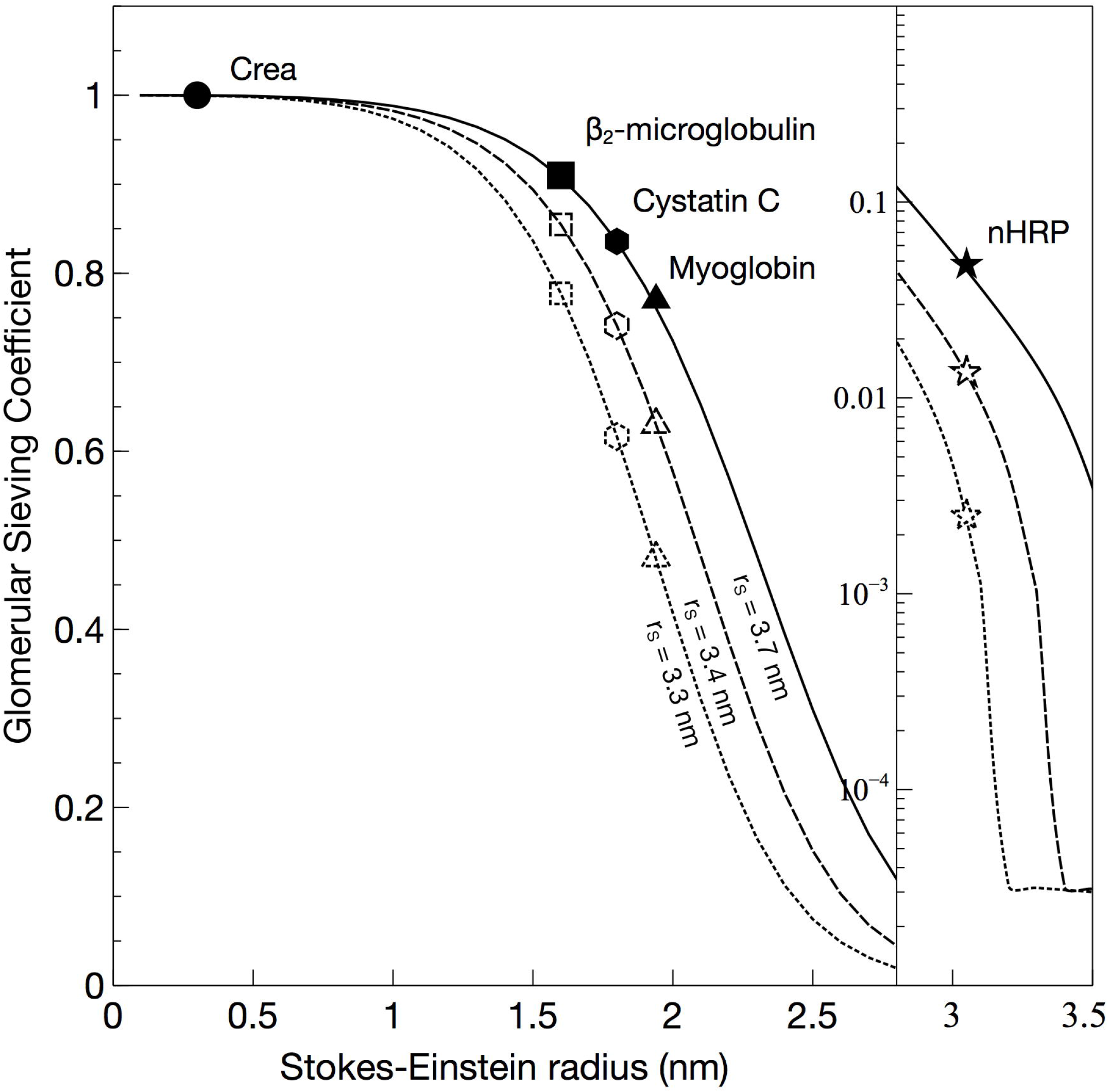
Glomerular sieving coefficients vs. SE-radius are for scenarios with shrunken pores, 3.4 nm (dashed line) and 3.2 nm (dotted line) compared to normal (3.7 nm; solid line).

**Figure 5.**
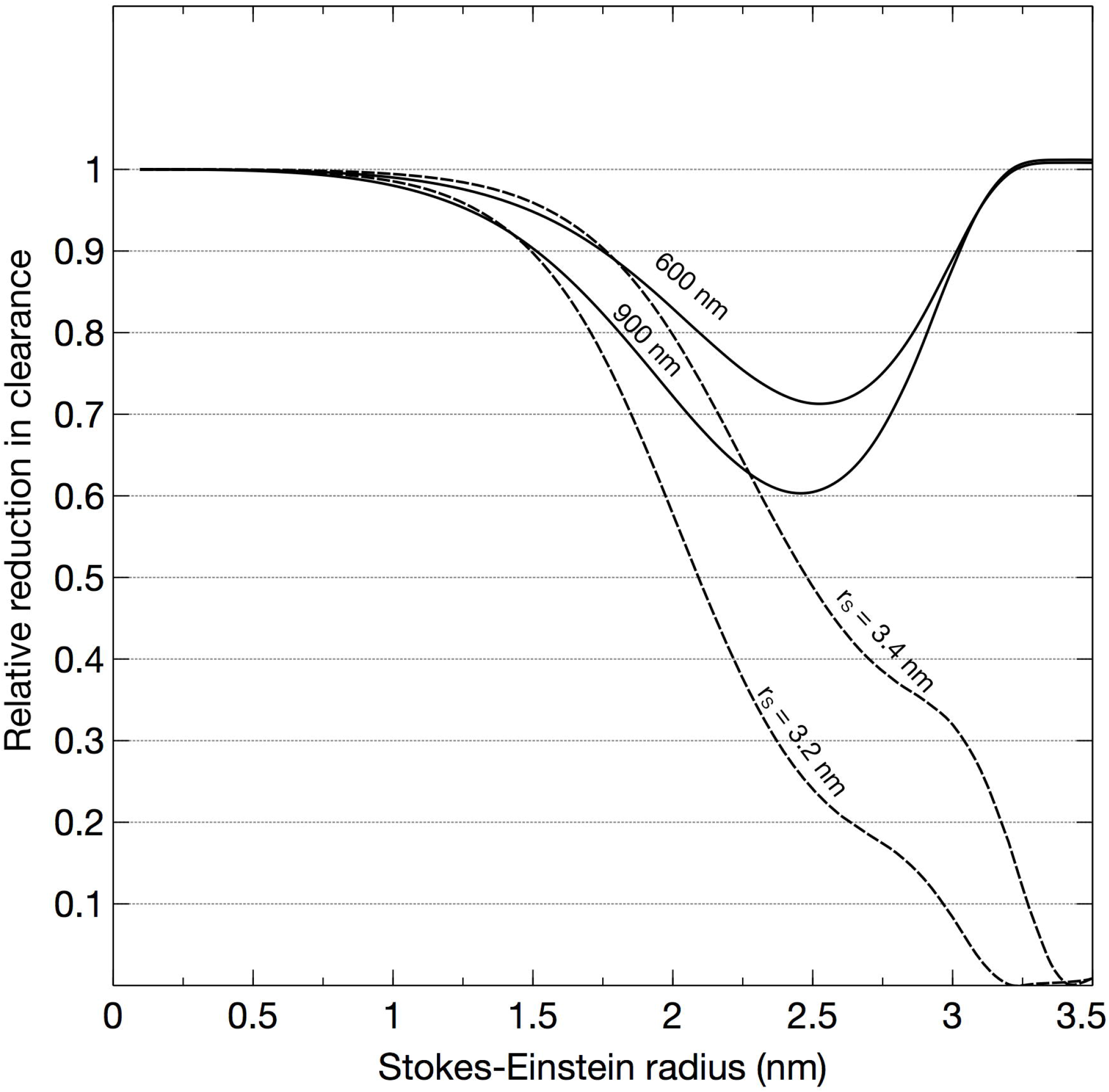
Pathological vs. healthy scenarios, either due to thickening of the glomerular filter (solid lines) to 600 nm or 900 nm, or shrinking of the functional pores (dashed lines) to 3.4 nm or 3.2 nm.

## Discussion

We here provide theoretical modeling and clinical data suggesting that the diffusive clearance of middle-molecular species such as cystatin C is reduced in DKD to a degree corresponding with the increment in the thickness of the glomerular basal membrane. We demonstrate a linear relationship between the ratio eGFR_cystatin C_/eGFR_creatinine_ and basal membrane thickness. According to theory, during conditions of glomerular vascular wall thickening, there should be a measurable difference in the eGFR measured using cystatin C and the actual GFR (e.g. as measured by iohexol) under the assumption that their plasma concentrations are determined chiefly by glomerular filtration. In line with our present results, a decreased diffusional clearance of mid-sized molecules has been found in experimental and clinical studies of diabetic kidney disease ^12, 32, 33^. There were however considerable variations in the eGFR_cystatin C_/eGFR_creatinine_ ratio in our data indicating that factors other than glomerular filtration may be involved. It is well-known that the steady-state plasma concentration of creatinine is affected by a number of other factors ^34^ which limits its use as a reliable GFR-marker. Similarly, cystatin C concentrations are affected by conditions such as high-dose corticosteroid medication ^35^. A limitation in the current study is therefore that it cannot be excluded that factors other than glomerular filtration affected the plasma levels of cystatin C and/or creatinine. A low muscle mass could give a higher eGFR_creatinine_ and thus lower eGFR_cystatin C_/eGFR_creatinine_ ratio. Regarding the effect of muscle mass on shrunken pore syndrome we could not find a correlation between BMI and eGFR_cystatin C_/eGFR_creatinine_ ratio. It has been described increased levels of cystatin C after high-dose corticosteroids which would give a low eGFR_cystatin C_/eGFR_creatinine_ ratio. In our cohort only three patients with MCD were treated with at least 30 mg prednisolon at the time of biopsy and no patients with DKD, which rules out an effect of steroids on our results. Recently the shrunken pore syndrome was reported which is characterized by a low ratio eGFR_cystatin C_/eGFR_creatinine_, and we wanted to study this ratio in correlation to the thickness of the basal membrane. However, the question of whether shrunken pore syndrome is the result of actual shrunken pores or vascular wall hypertrophy cannot be determined from the current data. Theoretically, shrinking of pores has more dramatic effects on the renal clearance of larger middle-molecules between 3.0 - 3.5 nm compared to small molecules, like creatinine and urea. Interestingly, many important plasma proteins, enzymes and peptide hormones are in this size range (e.g. cardiac Troponin T, TSH, AST, ALT and albumin) but many of them are catabolized in other tissues and regulated via other mechanisms making it unclear what effect shrunken pores would have on their plasma concentrations. It is well established that GFR is an important determinant of plasma cardiac troponin T (cTnT) levels ^36, 37^. According to our calculations, plasma cTnT levels should be several fold higher in a patient with shrunken pore syndrome (∼20 times higher for the 3.2 nm pore size), if shrunken pore syndrome is caused by an actual “pore shrinkage”.

The bi-selective nature of the GFB is well described in the literature ^30^. For the renal glomerular microcirculation, the first theoretical description was introduced by Deen et al in 1985 ^29^ using a model consisting of two distinct pathways for solute transport. Thus far, the most “accurate” theoretical description of the GFB is similar to that of a highly size-selective mechanical cross-flow filter, meaning that most of the blood flow through the glomerular capillaries travels in a direction tangentially across the capillary wall rather than entering the renal filter. Also, the red blood cells travel through the capillary in a compressed “cell-by-cell fashion” and thus constantly comb the filter surface. Both of these mechanisms, tangential solvent flux and continuous combing of the endothelial glycocalyx by RBCs, may counteract clogging of the renal filter. However, inevitably, some solutes must enter the capillary wall and, thus, must be dealt with via other mechanisms. Podocytes have been shown to exhibit endocytosis (and transcytosis) of albumin which will contribute to the clearance of albumin from the glomerular filter ^38^.

To understand why only solutes between 1.0 – 3.0 nm are affected by a thicker GFB one must first understand the concept of diffusion capacity, the proportionality coefficient in Fick’s equation

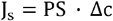

Here PS is the diffusion capacity in mL/min, J_s_ is the diffusive solute flux in mmol/min and Δc is the concentration gradient (Plasma water concentration – Bowman’s space concentration) in mmol/mL. From the equation above it can be seen that the diffusion capacity represents the maximal possible absolute diffusive clearance of a solute, being approached as the concentration on one side of the barrier is approaching zero. Mathematically, PS is the product of the diffusion coefficient D and the effective surface area A of the membrane divided by the membrane thickness Δx (i.e. PS = D × A / Δx), which means that PS will be reduced if the thickness, or diffusion length, Δx, is increased. Due to their small size, solutes like creatinine, urea, glucose *et cetera* have a great capacity to be transported via diffusion over the GFB since they have a very large diffusion capacity. Yet, diffusion is a process that requires a concentration gradient; and since small solutes < 1.0 nm are not sieved at all by the GFB Δc = 0 they will be transported entirely via convection and their clearance will equal GFR. For mid-sized solutes 1.0 – 3.0 nm there is a small but important concentration difference across the renal filter, leading to a considerable part of their clearance being due to diffusion due to the fact that the diffusion capacity is still very high (>> higher than GFR) for these solutes. With increasing solute sizes > 3.0 nm the diffusion capacity rapidly drops to low values (<< lower than GFR), and the diffusive clearance becomes negligible.

### Strengths

A validated registry for renal biopsy in which GFR was estimated from both creatinine and cystatin C is a strength of this study. The ultrastructural morphometry of the thickness of the GBM was evaluated by an experienced pathologist on enlarged electron micrographs as the arithmetic mean of 20 orthogonal intercepts across the GBM.

### Limitations

A major limitation is the low number of patients in both groups. However, the differences between the two groups, regarding eGFR_cystatin C_/eGFR_creatinine_ ratio and GBM thickness, were statistically significant.

*In conclusion* DKD develops insidiously over several years, well before any clinical manifestations (e.g. declining GFR and microproteinuria) are evident, and once microproteinuria is present, structural lesions are often considerably advanced. Caramori *et al* analyzed 94 biopsies in normoalbuminuric patients, and found that GBM thickness was the only structural parameter that predicted progression to diabetic nephropathy. Our data show that basal membrane thickness is associated with the clearance of freely filtered molecules in relation to their molecular weight obvious from a reduced ratio eGFR_cystatin C_/eGFR_creatinine_.

In light of our current findings, further research should explore the possibility that plasma levels of mid-sized molecules like cystatin C could act as bio-markers for detecting DKD, allowing early treatment. The ratio eGFR_cystatin C_/eGFR_creatinine_ may also be helpful in diagnosing other cases of CKD in early phases.

## Author contributions

CAÖ and AC designed the study and collected data. CAÖ performed mathematical modeling. CAÖ and AC analyzed the data. CAÖ made the figures. ML analyzed kidney biopsies. CAÖ, AG and AC drafted the manuscript. All authors contributed to, and approved the final version of the manuscript.

## Data Availability

All available data can be obtained from the corresponding author upon request.

## Acknowledgements

We dedicate this work to the memory of Professor Bengt Rippe (Gothenburg, 1950 – Lund, 2016), whose seminal work on the theoretical basis of fluid and solute transport across glomerular capillaries is now well recognized. The engaging and enthusiast personality of Bengt Rippe inspired a whole generation of physician-scientists. This work was funded by the medical faculty of Lund-Malmö (ALF-grant), the Swedish Kidney Foundation, Njurstiftelsen, Skåne University Hospital Research Fund and the Research and Development Council of Region Skåne, Sweden.

## Supplemental Material

This article contains the following supplemental material

Supplemental File 1: STARD-checklist

